# Tricuspid Valve Remodeling in a New Grading Scheme for Functional Tricuspid Regurgitation: A Three-Dimensional Echocardiography Study

**DOI:** 10.64898/2026.05.27.26354283

**Authors:** Yi Zhou, Yuji Xie, Xiaojun Yan, He Li, Mingxing Xie

## Abstract

**Background:** The specific 3D morphological substrates distinguishing the newly defined “massive” and “torrential” functional tricuspid regurgitation (FTR) phenotypes from standard severe disease remain under-characterized.

**Objectives:** This study investigates the 3D geometric changes of the tricuspid valve (TV) apparatus across the spectrum of FTR, specifically focusing on the structural definition of massive and torrential grades.

**Methods:** Three-dimensional (3D) transesophageal echocardiography (TEE) was performed in 322 patients with FTR secondary to left-sided heart disease. Patients were stratified into mild-moderate (n=166), severe (n=82), and massive-torrential (n=74) groups. TV geometry, including annular dimensions, leaflet tethering, and subvalvular apparatus, was quantified using 3D modeling software.

**Results:** Patients with massive-torrential TR were characterized by advanced age, female predominance, and atrial fibrillation (75%). 3D analysis demonstrated that massive-torrential TR represents a distinct phenotype defined by extreme annular circularization (ellipticity index ≈ 1.0) and planar flattening (*P* < 0.001). Furthermore, these patients exhibited a critical “leaflet-annulus uncoupling”, where compensatory leaflet growth (relative length < 80%) failed to match the massive annular dilation. Consequently, the regurgitant orifice in massive-torrential grades appeared highly complex, frequently manifesting as multiple irregular orifices.

**Conclusions:** Massive and torrential FTR are characterized by a unique geometric profile involving extreme annular circularization, severe leaflet tethering, and leaflet-annulus uncoupling. These morphological insights suggest that conventional repair strategies may be insufficient for these advanced phenotypes, highlighting the necessity for pre-procedural 3D TEE to guide device selection.

## Introduction

Functional tricuspid regurgitation (FTR) has historically been underappreciated, yet it represents a progressive pathology characterized by maladaptive geometric changes of the valvular apparatus in the absence of primary organic leaflet lesions ^1, 2^. While often initiated by left-sided heart disease or pulmonary hypertension, FTR can evolve into a self-perpetuating disease state driven by right atrial (RA) and right ventricular (RV) dilatation ^3, 4^.

Recently, the “massive” and “torrential” grades have been incorporated into the TR severity classification to address the “ceiling effect” of the conventional grading system and improve prognostic stratification in transcatheter intervention trials ^5, 6^. Despite the rapid expansion of transcatheter tricuspid valve interventions, the specific morphological substrates defining “massive” and “torrential” tricuspid regurgitation (TR) remain incompletely elucidated. Current therapeutic failures, particularly recurrent TR after annuloplasty ^7, 8^, are frequently attributed to to an incomplete understanding of complex 3D tethering mechanisms and unrecognized leaflet-to-annulus mismatch ^9–11^.

Consequently, this study aims to utilize 3D Transesophageal echocardiography (TEE) to comprehensively characterize the geometric phenotype of massive and torrential FTR ^12^. We hypothesized that these advanced grades constitute a distinct morphological entity characterized by “leaflet-annulus uncoupling”, a finding that would have direct implications for anatomical eligibility and device selection in transcatheter tricuspid valve replacement (TTVR) trials.

## Methods

### Patient Population

We retrospectively analyzed the records of 322 consecutive patients with FTR who underwent 3D TEE at Union Hospital between June 2020 and August 2022. The inclusion criterion was FTR associated with left-sided heart disease. Exclusion criteria were as follows: 1) primary tricuspid valve (TV) disease (e.g., rheumatic, prolapse, carcinoid); 2) cardiac implantable electronic device leads across the TV; 3) congenital heart disease; and 4) inadequate 3D TV visualization (Figure S1 and S2). The final population was classified into three groups based on TR severity: mild-moderate (n=166), severe (n=82), and massive-torrential (n=74).

### Echocardiographic Acquisition

Comprehensive 2D and 3D echocardiographic assessments were conducted using an EPIQ 7 ultrasound system (Philips Medical Systems, Andover, MA, USA) equipped with a fully sampled matrix-array transducer (X7-2t or X8-2t Live 3D TEE transducer) capable of high-volume rate display. 3D full-volume datasets or 3D zoom loops of the TV were acquired from the mid-oesophageal RV-focused inflow-outflow view. Views were optimized for depth and gain settings to ensure optimal temporal and spatial resolution ^13^. A multi-beat acquisition mode (5 beats) was employed for patients in sinus rhythm, whereas a single-beat mode was utilized for patients with atrial fibrillation (AF) to prevent stitching artifacts.

### 3D Image Analysis

Data analysis was performed offline using commercially available quantification software (QLAB ver. 10.5, Philips Medical Systems).

### Assessment of TR severity

TR severity was graded using the expanded scheme based on vena contracta width (VCW) and 3D vena contracta area (VCA) ^14^. The VCW was averaged from two orthogonal views^15^. For 3D VCA, the Color Doppler dataset was cropped using multiplanar reconstruction to manually trace the regurgitant orifice area at the narrowest neck of the jet (mid-systole) ^16^. Patients were stratified as: mild–moderate (VCW < 7 mm), severe (VCW 7∼13.9 mm; 3D VCA 75∼94 mm^2^), and massive-torrential (VCW ≥ 14 mm; 3D VCA ≥ 95 mm^2^) ^14^.

### Quantitative Assessment of Tricuspid Valve Geometry

Morphological analysis of the TV apparatus was conducted using the Mitral Valve Quantification (MVQ) software (Philips Medical Systems) adapted for the TV. The 3D volume rendering was oriented with the interatrial septum positioned inferiorly at the 6 o’clock position (Figure 2A) ^17^.

### Annular Parameters

The tricuspid annulus (TA) was delineated using 20 landmarks. The anteroposterior (A-P) and anterolateral-posteromedial (AL-PM) axes were defined using the commissure between the anterior (ATL) and septal (STL) leaflets as the anatomical reference. ^18^ Annular dimensions—including area, circumference, height, and ellipticity (AL-PM/A-P ratio)—were measured at mid-systole and mid-diastole. TA orientation was quantified as the angle between the left atrium and the vertical axis at the level of the interatrial septum (Figure 2B).

### Leaflet and Tethering Parameters

The 3D Quantification (3DQ) software was used to generate 2D planes perpendicular to each leaflet to the coaptation line of each leaflet to measure tethering angles in mid-systole (Figure 2C). The TV tenting volume was calculated by tracing the closed leaflets on contiguous long-axis planes. Leaflet length and thickness were measured, and the “relative leaflet length” (ratio of absolute leaflet length to A-P annular diameter) was calculated to evaluate leaflet-to-annulus mismatch.

### Subvalvular Apparatus Parameters

The lengths of the anterior and septal papillary chordae tendineae were quantified using the MVQ software (Figure S3).

### Statistical analysis

Statistical analysis was performed using IBM SPSS Statistics version 26.0 (SPSS, Inc., Chicago, IL, USA). Continuous variables were tested for normality and expressed as mean ± standard deviation for normally distributed data or median (interquartile range [IQR]) for non-normally distributed data. Categorical variables were presented as frequencies (percentages) and compared using the *Chi-square test* or *Fisher’s exact test*. Comparisons among the mild-moderate, severe, and massive-torrential groups were performed using *ANOVA* for continuous variables. All statistical tests were two-sided, and a *P* value < 0.05 was considered statistically significant.

## Results

### Patient Population

The study population selection is summarized in the flowchart in Figure 1. From an initial cohort of 483 patients with TR, 3D TV screening identified 80 patients (16.6%) with primary TR who were excluded. The remaining 403 patients (83.4%) were classified as having FTR. Within this group, the majority (79.9%, n=322) presented with FTR secondary to left-sided heart disease.

**Figure 1.**
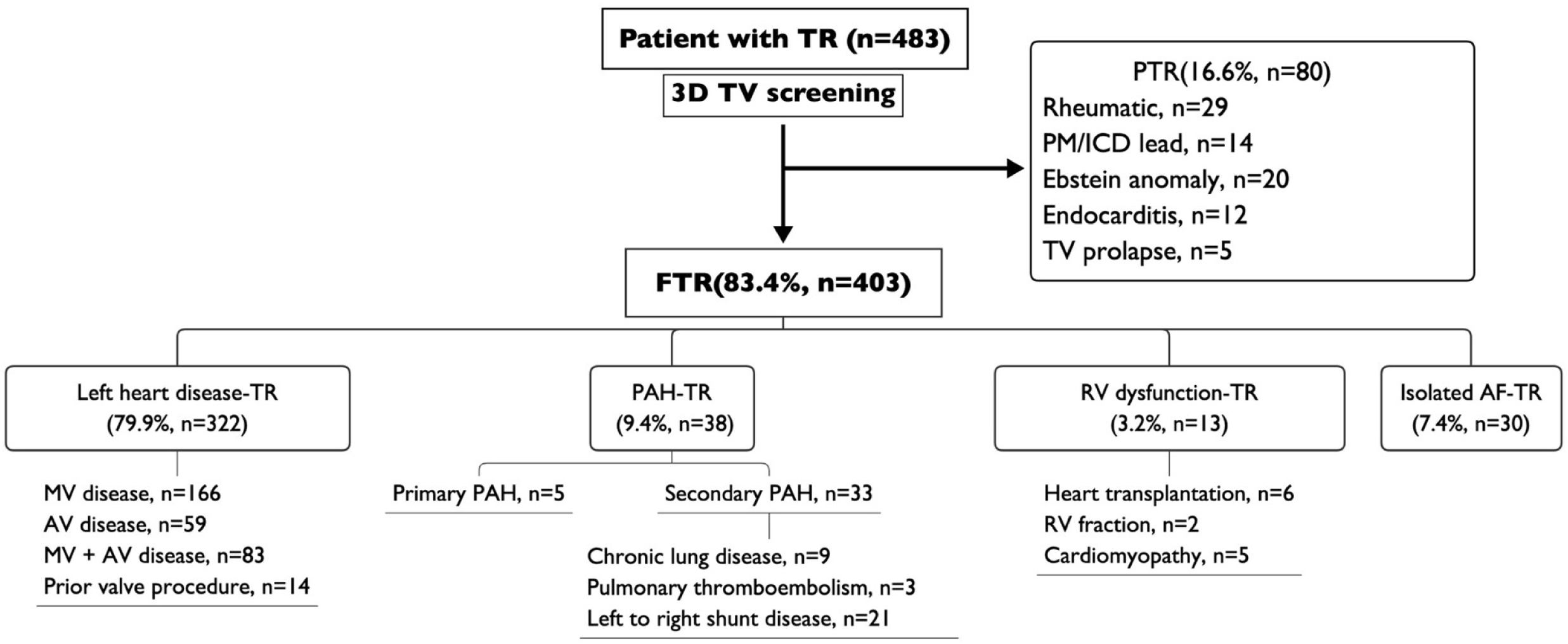
Distribution of various causes of TR assessed by 3D TEE. FTR was present in 322 patients (79.9%) due to left-side heart disease, 38 patients (9.4%) due to PAH, 30 patients 7.4%) due to isolated AF, and 13 patients (3.2%) due to RV dysfunction. TR indicates tricuspid regurgitation; TV, tricuspid valve; PTR, primary tricuspid regurgitation; PM, pacemaker; ICD, implantable cardioverter defibrillator; FTR, functional tricuspid regurgitation; PAH, pulmonary arterial hypertension; AV, aortic valve; MV, mitral valve; RV, right ventricular; AF, atrial fibrillation.

**Figure 2.**
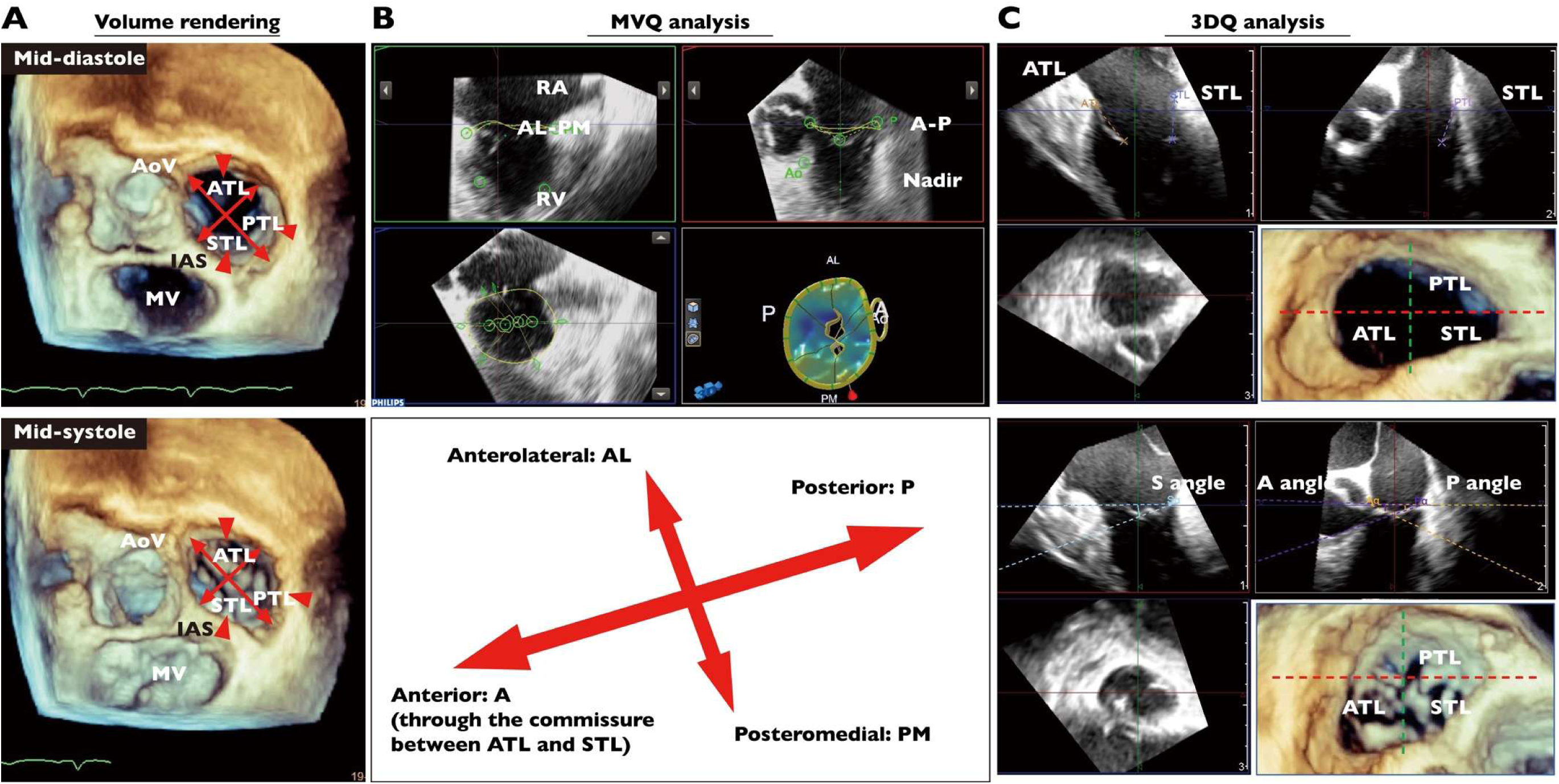
Three-dimensional TEE-based TV data acquisition and analysis by mitral valve quantification (MVQ) software and 3D quantification (3DQ) software. A, 3D volume rendering. The TV on 3D TEE as viewed from the RA perspective in mid-diastole and mid-systole. All 3 commissures of the leaflets were identified (red arrowheads). B, MVQ. A-P and AL-PM directions were determined using the commissure between the anterior and septal leaflets as the anterior aspect (arrows). C, 3DQ. The 3 long-axis planes, which perpendicularly crossed the middle of each tricuspid leaflet, were generated carefully using guidance on the short-axis image of the TV. On each of the 3 long-axis planes, the tethering angles of the leaflets were measured on a mid-systole frame. The leaflet lengths of the ATL, PTL, and STL were measured on mid-diastole frames, and (Aα, Pα and Sα) the closure lengths were measured on mid-systole frames. AoV indicates aortic valve; ATL, anterior tricuspid leaflet; IAS, interatrial septum; MV, mitral valve; PTL, posterior tricuspid leaflet; STL, septal tricuspid leaflet; RV, right ventricle; AL anterolateral, and PM, posteromedial.

The massive–torrential TR cohort (n=74) exhibited a distinct clinical profile compared to the mild–moderate (n=166) and severe (n=82) groups. These patients were significantly older (55 ± 15 years) and predominantly female (59.5% vs. 34.8% in mild–moderate; *P* < 0.001). The prevalence of AF increased markedly with disease severity, reaching 75.3% in the massive–torrential group compared to 27.3% in the mild–moderate group (*P* < 0.001). Advanced TR was also associated with a higher burden of comorbidities, including prior stroke, renal impairment (lower estimated glomerular filtration rate), and thrombocytopenia (*P* < 0.05). Functional impairment was pronounced in the massive–torrential group, characterized by elevated B-type natriuretic peptide levels and advanced NYHA class (90.5% in class III). Notably, ventilation dysfunction was significantly more prevalent in massive–torrential patients (*P* < 0.001) (Table 1).

**TABLE 1.**
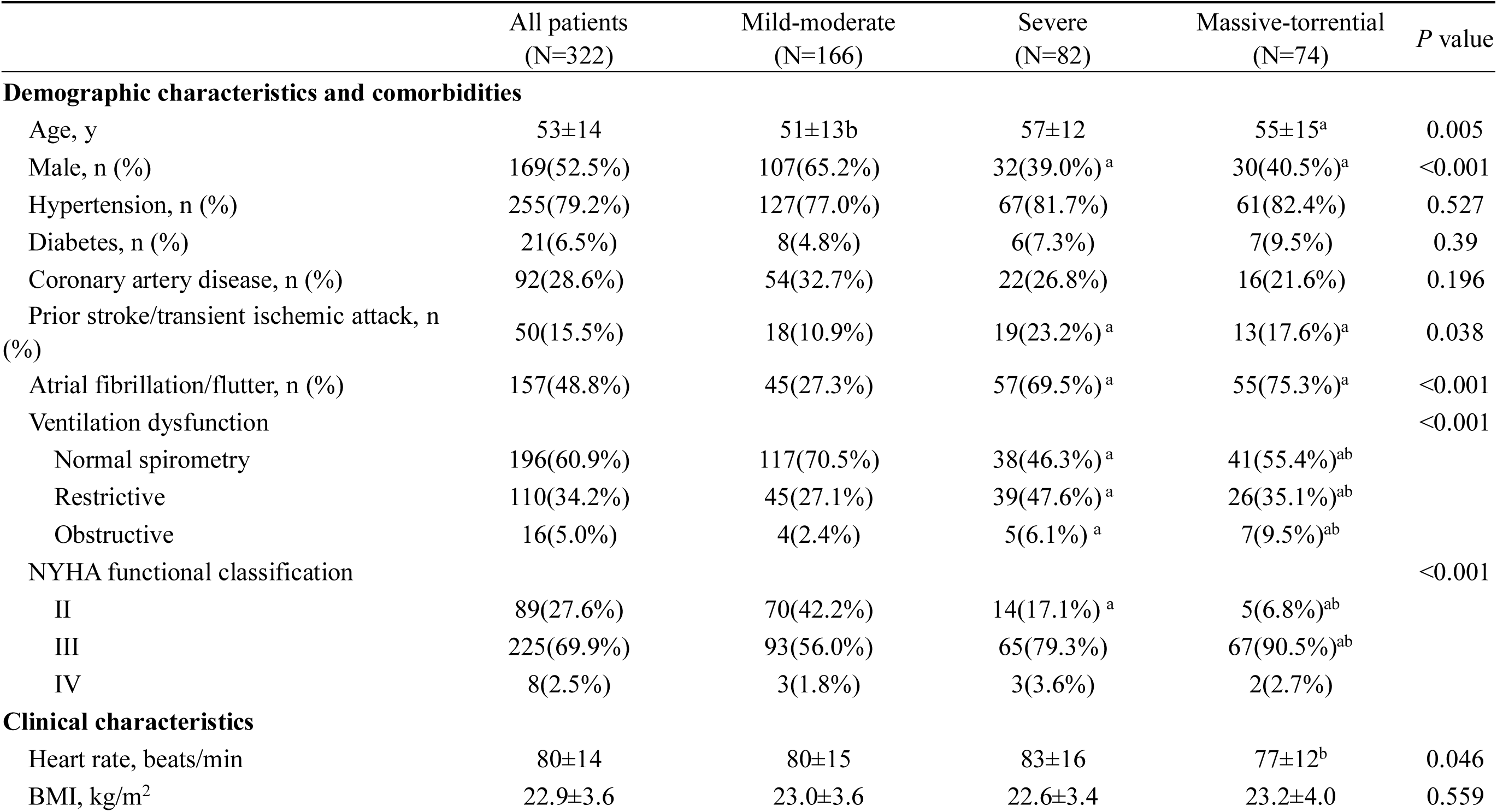
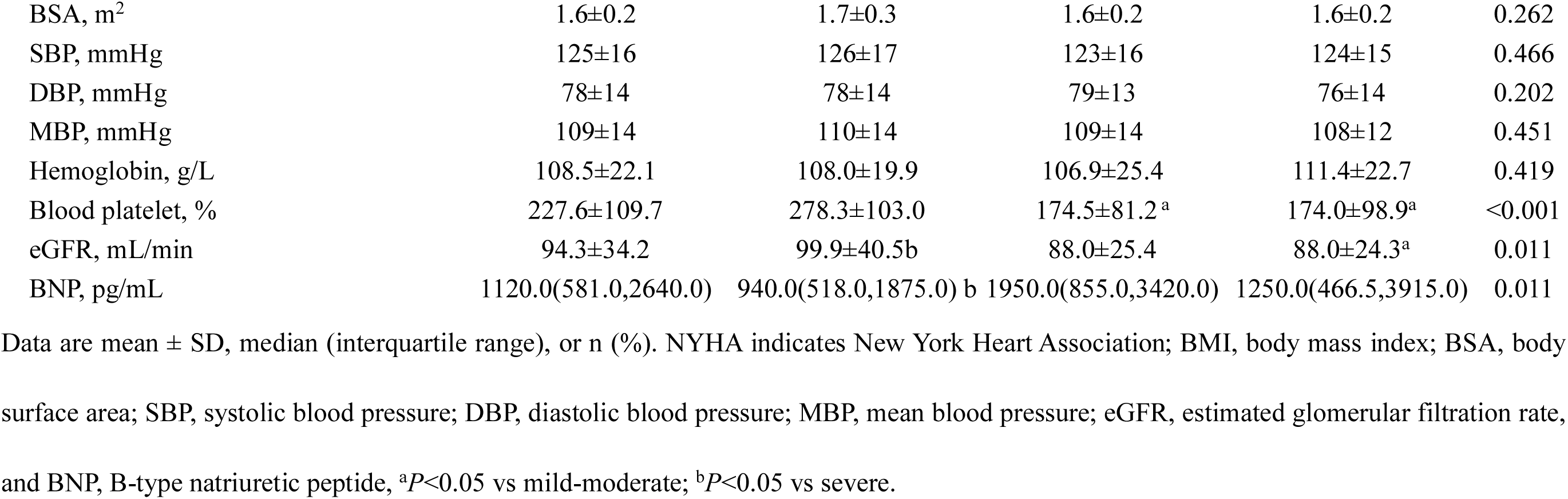
Baseline characteristics of the study population (n=332)

|  | All patients<br>(N=322) | Mild-moderate<br>(N=166) | Severe<br>(N=82) | Massive-torrential<br>(N=74) | *P* value |
| --- | --- | --- | --- | --- | --- |
| **Demographic characteristics and comorbidities** |  |  |  |  |  |
| Age, y | 53±14 | 51±13b | 57±12 | 55±15<sup>a</sup> | 0.005 |
| Male, n (%) | 169(52.5%) | 107(65.2%) | 32(39.0%)<sup>a</sup> | 30(40.5%)<sup>a</sup> | <0.001 |
| Hypertension, n (%) | 255(79.2%) | 127(77.0%) | 67(81.7%) | 61(82.4%) | 0.527 |
| Diabetes, n (%) | 21(6.5%) | 8(4.8%) | 6(7.3%) | 7(9.5%) | 0.39 |
| Coronary artery disease, n (%) | 92(28.6%) | 54(32.7%) | 22(26.8%) | 16(21.6%) | 0.196 |
| Prior stroke/transient ischemic attack, n (%) | 50(15.5%) | 18(10.9%) | 19(23.2%)<sup>a</sup> | 13(17.6%)<sup>a</sup> | 0.038 |
| Atrial fibrillation/flutter, n (%) | 157(48.8%) | 45(27.3%) | 57(69.5%)<sup>a</sup> | 55(75.3%)<sup>a</sup> | <0.001 |
| Ventilation dysfunction |  |  |  |  | <0.001 |
| Normal spirometry | 196(60.9%) | 117(70.5%) | 38(46.3%)<sup>a</sup> | 41(55.4%)<sup>ab</sup> |  |
| Restrictive | 110(34.2%) | 45(27.1%) | 39(47.6%)<sup>a</sup> | 26(35.1%)<sup>ab</sup> |  |
| Obstructive | 16(5.0%) | 4(2.4%) | 5(6.1%)<sup>a</sup> | 7(9.5%)<sup>ab</sup> |  |
| NYHA functional classification |  |  |  |  | <0.001 |
| II | 89(27.6%) | 70(42.2%) | 14(17.1%)<sup>a</sup> | 5(6.8%)<sup>ab</sup> |  |
| III | 225(69.9%) | 93(56.0%) | 65(79.3%) | 67(90.5%)<sup>ab</sup> |  |
| IV | 8(2.5%) | 3(1.8%) | 3(3.6%) | 2(2.7%) |  |
| **Clinical characteristics** |  |  |  |  |  |
| Heart rate, beats/min | 80±14 | 80±15 | 83±16 | 77±12<sup>b</sup> | 0.046 |
| BMI, kg/m<sup>2</sup> | 22.9±3.6 | 23.0±3.6 | 22.6±3.4 | 23.2±4.0 | 0.559 |
| BSA, m<sup>2</sup> | 1.6±0.2 | 1.7±0.3 | 1.6±0.2 | 1.6±0.2 | 0.262 |
| SBP, mmHg | 125±16 | 126±17 | 123±16 | 124±15 | 0.466 |
| DBP, mmHg | 78±14 | 78±14 | 79±13 | 76±14 | 0.202 |
| MBP, mmHg | 109±14 | 110±14 | 109±14 | 108±12 | 0.451 |
| Hemoglobin, g/L | 108.5±22.1 | 108.0±19.9 | 106.9±25.4 | 111.4±22.7 | 0.419 |
| Blood platelet, % | 227.6±109.7 | 278.3±103.0 | 174.5±81.2 <sup>a</sup> | 174.0±98.9<sup>a</sup> | <0.001 |
| eGFR, mL/min | 94.3±34.2 | 99.9±40.5<sup>b</sup> | 88.0±25.4 | 88.0±24.3<sup>a</sup> | 0.011 |
| BNP, pg/mL | 1120.0(581.0,2640.0) | 940.0(518.0,1875.0) <sup>b</sup> | 1950.0(855.0,3420.0) | 1250.0(466.5,3915.0) | 0.011 |

Data are mean ± SD, median (interquartile range), or n (%). NYHA indicates New York Heart Association; BMI, body mass index; BSA, body surface area; SBP, systolic blood pressure; DBP, diastolic blood pressure; MBP, mean blood pressure; eGFR, estimated glomerular filtration rate, and BNP, B-type natriuretic peptide, <sup>a</sup>*P*<0.05 vs mild-moderate; <sup>b</sup>*P*<0.05 vs severe.

### Echocardiographic characteristics

Conventional echocardiography demonstrated significant cardiac remodeling proportional to TR severity. Left atrium (LA) and left ventricular (LV) diameters were significantly larger in severe and massive-torrential patients compared to the mild-moderate group (*P* < 0.001). Right-sided remodeling was particularly pronounced; RA diameter increased from 3.9 ± 0.6 cm in the mild-moderate group to 5.5 ± 1.2 cm in the massive-torrential group (*P* < 0.001), with a similar increase in RV diameter (3.6 ± 0.5 cm vs. 4.6 ± 0.8 cm; *P* < 0.001). Hemodynamically, pulmonary artery systolic pressure rose significantly with TR severity, averaging 46.3 ± 17.6 mmHg in the massive-torrential group compared to 34.4 ± 18.6 mmHg in the mild-moderate group (*P* < 0.001). The prevalence of concomitant severe mitral, aortic, and pulmonary regurgitation was also significantly higher in the advanced TR groups. Additionally, the inferior vena cava diameter was significantly dilated with a reduced collapse rate in the massive-torrential group (*P* < 0.001) (Table 2).

**TABLE 2.**
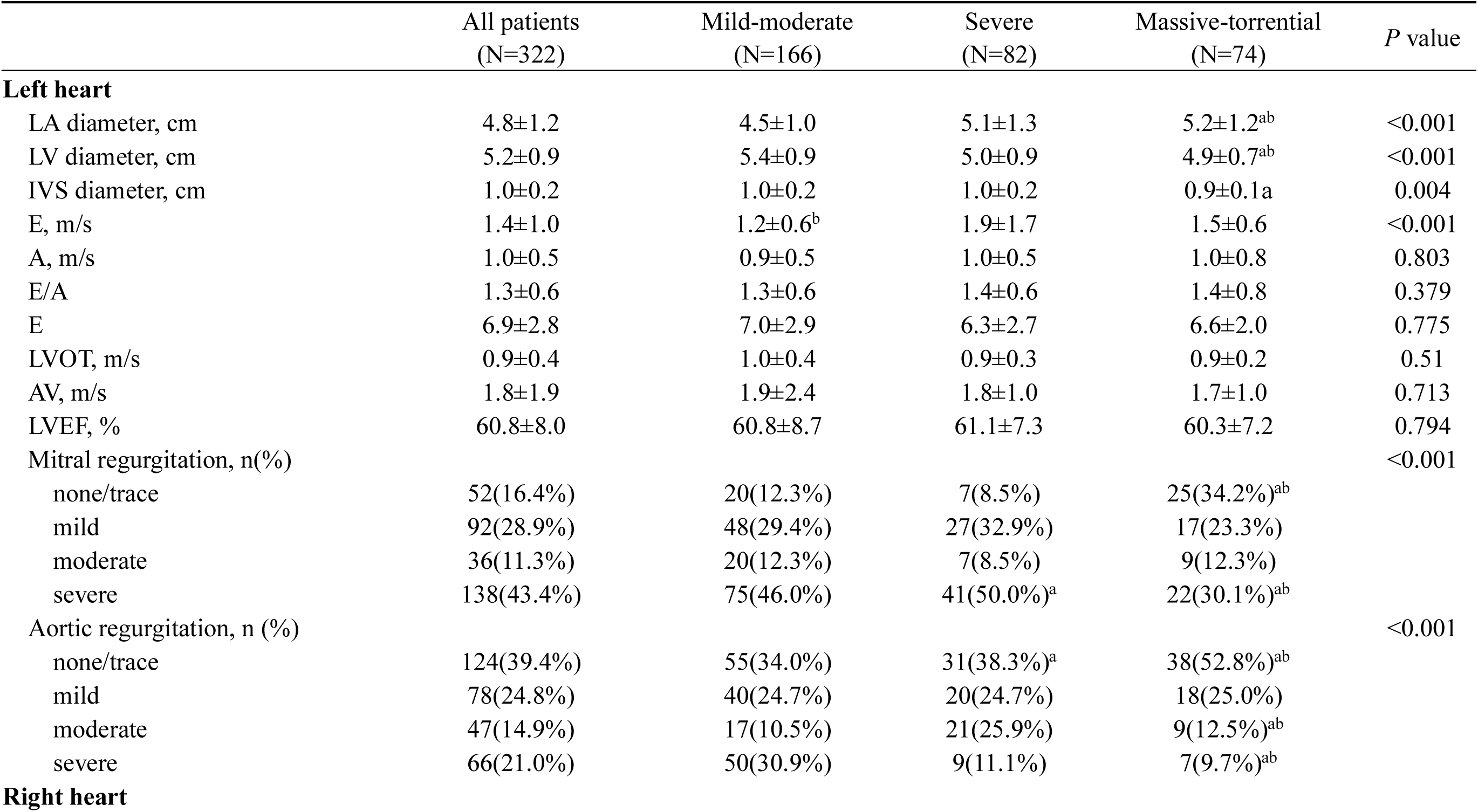
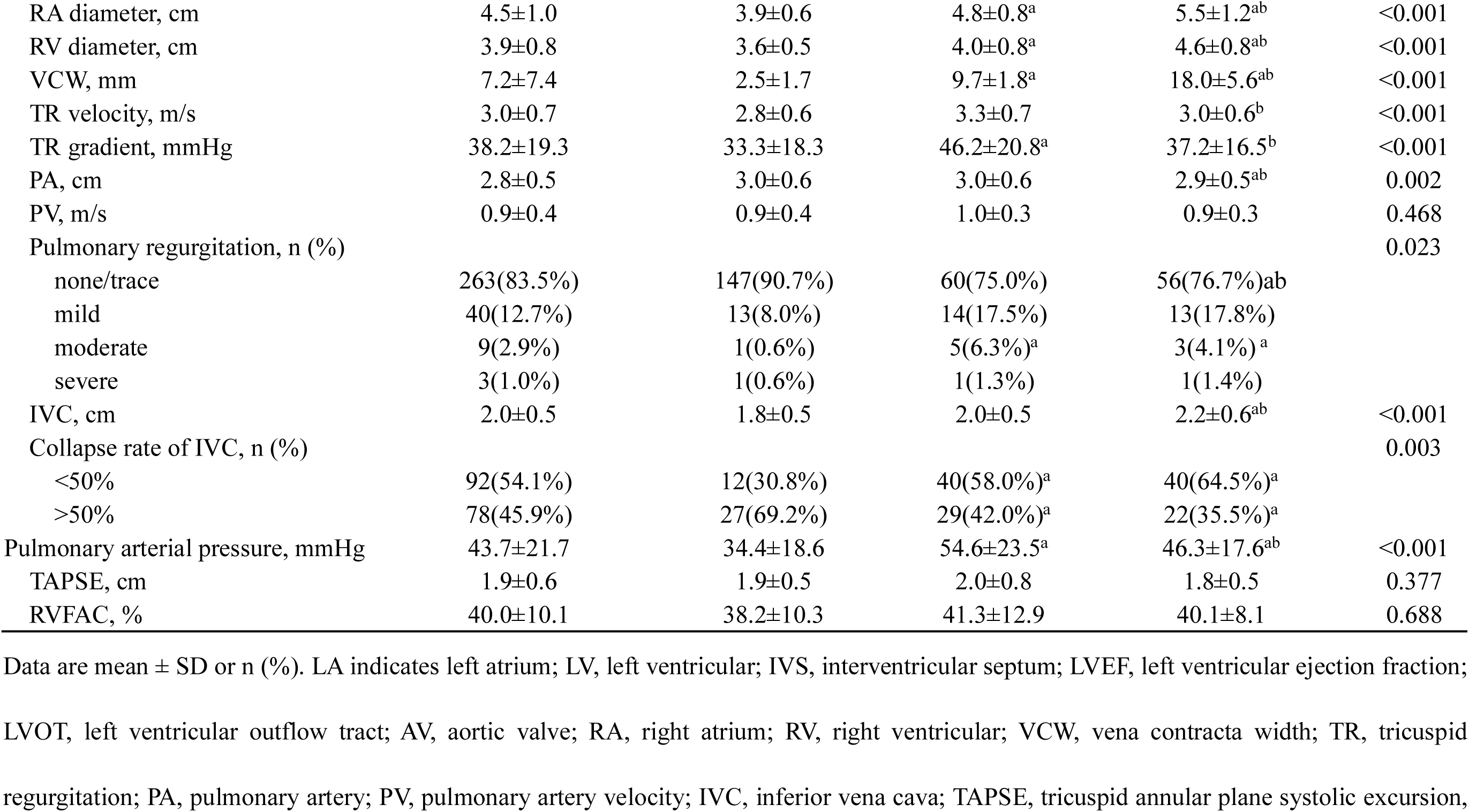
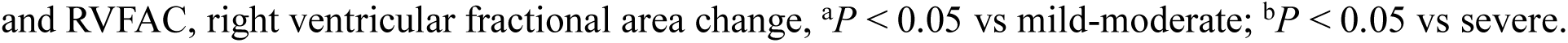
Conventional echocardiographic parameters(n=332)

|  | All patients<br>(N=322) | Mild-moderate<br>(N=166) | Severe<br>(N=82) | Massive-torrential<br>(N=74) | *P* value |
| --- | --- | --- | --- | --- | --- |
| **Left heart** |  |  |  |  |  |
| LA diameter, cm | 4.8±1.2 | 4.5±1.0 | 5.1±1.3 | 5.2±1.2<sup>ab</sup> | <0.001 |
| LV diameter, cm | 5.2±0.9 | 5.4±0.9 | 5.0±0.9 | 4.9±0.7<sup>ab</sup> | <0.001 |
| IVS diameter, cm | 1.0±0.2 | 1.0±0.2 | 1.0±0.2 | 0.9±0.1<sup>a</sup> | 0.004 |
| E, m/s | 1.4±1.0 | 1.2±0.6<sup>b</sup> | 1.9±1.7 | 1.5±0.6 | <0.001 |
| A, m/s | 1.0±0.5 | 0.9±0.5 | 1.0±0.5 | 1.0±0.8 | 0.803 |
| E/A | 1.3±0.6 | 1.3±0.6 | 1.4±0.6 | 1.4±0.8 | 0.379 |
| E | 6.9±2.8 | 7.0±2.9 | 6.3±2.7 | 6.6±2.0 | 0.775 |
| LVOT, m/s | 0.9±0.4 | 1.0±0.4 | 0.9±0.3 | 0.9±0.2 | 0.51 |
| AV, m/s | 1.8±1.9 | 1.9±2.4 | 1.8±1.0 | 1.7±1.0 | 0.713 |
| LVEF, % | 60.8±8.0 | 60.8±8.7 | 61.1±7.3 | 60.3±7.2 | 0.794 |
| Mitral regurgitation, n(%) |  |  |  |  | <0.001 |
| none/trace | 52(16.4%) | 20(12.3%) | 7(8.5%) | 25(34.2%)<sup>ab</sup> |  |
| mild | 92(28.9%) | 48(29.4%) | 27(32.9%) | 17(23.3%) |  |
| moderate | 36(11.3%) | 20(12.3%) | 7(8.5%) | 9(12.3%) |  |
| severe | 138(43.4%) | 75(46.0%) | 41(50.0%)<sup>a</sup> | 22(30.1%)<sup>ab</sup> |  |
| Aortic regurgitation, n (%) |  |  |  |  | <0.001 |
| none/trace | 124(39.4%) | 55(34.0%) | 31(38.3%)<sup>a</sup> | 38(52.8%)<sup>ab</sup> |  |
| mild | 78(24.8%) | 40(24.7%) | 20(24.7%) | 18(25.0%) |  |
| moderate | 47(14.9%) | 17(10.5%) | 21(25.9%) | 9(12.5%)<sup>ab</sup> |  |
| severe | 66(21.0%) | 50(30.9%) | 9(11.1%) | 7(9.7%)<sup>ab</sup> |  |
| **Right heart** |  |  |  |  |  |
| RA diameter, cm | 4.5±1.0 | 3.9±0.6 | 4.8±0.8<sup>a</sup> | 5.5±1.2<sup>ab</sup> | <0.001 |
| RV diameter, cm | 3.9±0.8 | 3.6±0.5 | 4.0±0.8<sup>a</sup> | 4.6±0.8<sup>ab</sup> | <0.001 |
| VCW, mm | 7.2±7.4 | 2.5±1.7 | 9.7±1.8<sup>a</sup> | 18.0±5.6<sup>ab</sup> | <0.001 |
| TR velocity, m/s | 3.0±0.7 | 2.8±0.6 | 3.3±0.7 | 3.0±0.6<sup>b</sup> | <0.001 |
| TR gradient, mmHg | 38.2±19.3 | 33.3±18.3 | 46.2±20.8<sup>a</sup> | 37.2±16.5<sup>b</sup> | <0.001 |
| PA, cm | 2.8±0.5 | 3.0±0.6 | 3.0±0.6 | 2.9±0.5<sup>ab</sup> | 0.002 |
| PV, m/s | 0.9±0.4 | 0.9±0.4 | 1.0±0.3 | 0.9±0.3 | 0.468 |
| Pulmonary regurgitation, n (%) |  |  |  |  | 0.023 |
| none/trace | 263(83.5%) | 147(90.7%) | 60(75.0%) | 56(76.7%)<sup>ab</sup> |  |
| mild | 40(12.7%) | 13(8.0%) | 14(17.5%) | 13(17.8%) |  |
| moderate | 9(2.9%) | 1(0.6%) | 5(6.3%)<sup>a</sup> | 3(4.1%)<sup>a</sup> |  |
| severe | 3(1.0%) | 1(0.6%) | 1(1.3%) | 1(1.4%) |  |
| IVC, cm | 2.0±0.5 | 1.8±0.5 | 2.0±0.5 | 2.2±0.6<sup>ab</sup> | <0.001 |
| Collapse rate of IVC, n (%) |  |  |  |  | 0.003 |
| <50% | 92(54.1%) | 12(30.8%) | 40(58.0%)<sup>a</sup> | 40(64.5%)<sup>a</sup> |  |
| >50% | 78(45.9%) | 27(69.2%) | 29(42.0%)<sup>a</sup> | 22(35.5%)<sup>a</sup> |  |
| Pulmonary arterial pressure, mmHg | 43.7±21.7 | 34.4±18.6 | 54.6±23.5<sup>a</sup> | 46.3±17.6<sup>ab</sup> | <0.001 |
| TAPSE, cm | 1.9±0.6 | 1.9±0.5 | 2.0±0.8 | 1.8±0.5 | 0.377 |
| RVFAC, % | 40.0±10.1 | 38.2±10.3 | 41.3±12.9 | 40.1±8.1 | 0.688 |

Data are mean ± SD or n (%). LA indicates left atrium; LV, left ventricular; IVS, interventricular septum; LVEF, left ventricular ejection fraction; LVOT, left ventricular outflow tract; AV, aortic valve; RA, right atrium; RV, right ventricular; VCW, vena contracta width; TR, tricuspid regurgitation; PA, pulmonary artery; PV, pulmonary artery velocity; IVC, inferior vena cava; TAPSE, tricuspid annular plane systolic excursion,
and RVFAC, right ventricular fractional area change, <sup>a</sup> $P < 0.05$ vs mild-moderate; <sup>b</sup> $P < 0.05$ vs severe.

### 3D Morphological Remodeling of the TV

3D TEE analysis demonstrated a continuum of geometric distortion in the tricuspid apparatus, with the most extreme changes observed in the massive-torrential cohort (Table 3).

**TABLE 3.** Three-Dimensional TV Cohort(n=332)

|  | All patients<br>(N=322) | Mild-moderate<br>(N=166) | Severe<br>N=82) | Massive-torrential<br>(N=74) | *P* value |
| --- | --- | --- | --- | --- | --- |
| 3D VCA, mm<sup>2</sup> | 47.3±46.1 | 12.7±17.2 | 76.5±18.4<sup>a</sup> | 109.1±21.7<sup>ab</sup> | <0.001 |
| Atrial fibrillation/flutter, n (%) | 157(48.8%) | 45(27.3%) | 57(69.5%)<sup>a</sup> | 55(75.3%)<sup>a</sup> | <0.001 |
| **Tricuspid annular parameters** |  |  |  |  |  |
| Annular diameter (AL-PM), mm | 40.1±6.6 | 39.0±6.1 | 39.3±5.8 | 43.4±7.3<sup>a</sup> | <0.001 |
| Annular diameter(A-P), mm | 37.1±8.1 | 33.9±6.0 | 37.3±5.9<sup>a</sup> | 44.7±8.8<sup>ab</sup> | <0.001 |
| Annular height, mm | 5.1±1.8 | 5.5±1.8 | 4.6±1.8 | 4.6±1.5<sup>ab</sup> | 0.003 |
| 3D annular circumference, mm | 130.2±19.1 | 124.8±14.4 | 128.3±15.0 | 144.6±24.0<sup>ab</sup> | <0.001 |
| 3D annular area, mm<sup>2</sup> | 1274.5±421.9 | 1131.5±261.6 | 1238.5±296.2 | 1642.2±571.3<sup>ab</sup> | <0.001 |
| Ellipticity (AL-PM/A-P), % | 1.1±0.2 | 1.2±0.3 | 1.1±0.2<sup>a</sup> | 1.0±0.1<sup>a</sup> | <0.001 |
| Ao-tricuspid angle, ° | 108.4±15.5 | 108.2±15.4 | 106.2±15.6 | 110.5±15.9 | 0.468 |
| TA orientation, ° | 59.0±23.8 | 40.9±6.2 | 87.7±11.8<sup>a</sup> | 84.7±11.6<sup>a</sup> | <0.001 |
| **Tricuspid leaflet parameters** |  |  |  |  |  |
| 3D TV leaflet area, mm<sup>2</sup> | 1459.2±536.4 | 1290.4±375.0 | 1408.2±385.0 | 1899.2±697.7<sup>ab</sup> | <0.001 |
| Tenting volume, mL | 2.0±1.7 | 1.5±1.2 | 2.0±1.3 | 3.2±2.3<sup>ab</sup> | <0.001 |
| Tenting height, mm | 6.7±3.0 | 6.0±2.8 | 7.4±2.9<sup>a</sup> | 8.0±2.9<sup>a</sup> | <0.001 |
| Averaged tethering angle, ° | 24.2±11.9 | 15.2±4.4 | 28.0±2.2<sup>a</sup> | 42.5±3.0<sup>a</sup> | <0.001 |
| ATL angle, ° | 20.6±12.0 | 11.6±4.7 | 24.4±2.3<sup>a</sup> | 38.8±3.4<sup>ab</sup> | <0.001 |
| PTL angle, ° | 25.6±10.4 | 18.1±4.5 | 28.1±2.9<sup>a</sup> | 41.6±3.0<sup>ab</sup> | <0.001 |
| STL angle, ° | 26.3±13.4 | 15.8±4.7 | 31.4±2.8<sup>a</sup> | 47.2±3.0<sup>ab</sup> | <0.001 |
| Total leaflet relative length, % | 135.9% ± 48.1 | 167.9% ± 39.5 | 118.3% ± 29.8<sup>a</sup> | 79.7% ± 28.0<sup>ab</sup> | <0.001 |
| ATL relative length, % | 57.1% ± 19.3 | 69.2% ± 16.5 | 52.4% ± 12.3<sup>a</sup> | 35.3% ± 12.5<sup>ab</sup> | <0.001 |
| PTL relative length, % | 44.9% ± 16.6 | 56.2% ± 13.5 | 38.4% ± 9.5<sup>a</sup> | 25.3% ± 9.5<sup>ab</sup> | <0.001 |
| STL relative length, % | 34.7% ± 13.0 | 44.1% ± 10.1 | 29.0% ± 8.0<sup>a</sup> | 19.1% ± 6.6<sup>ab</sup> | <0.001 |
| **Subvalvular apparatus parameters** |  |  |  |  |  |
| Anterior papillary chordae tendinea length, mm | 28.6±9.4 | 26.1±10.0 | 29.4±6.2 | 34.0±7.4<sup>ab</sup> | <0.001 |
| Posterior papillary chordae tendinea length, mm | 32.7±9.6 | 31.3±10.3 | 32.4±6.2 | 36.4±9.1<sup>a</sup> | 0.009 |
| Height H, mm | 25.3±3.9 | 24.3±2.7 | 25.5±2.7<sup>a</sup> | 27.6±5.7<sup>ab</sup> | <0.001 |
| S<sub>ΔABC</sub>, mm<sup>2</sup> | 405.3(334.9,405.3) | 335.8(308.6,395.9) | 453.9(406.1,481.6) | 583.5(485.6,709.8)<sup>ab</sup> | <0.001 |

Data are mean ± SD or n (%). VCA indicates vena contracta area; A indicates anterior; P, posterior; S, septum; AL anterolateral; PM, posteromedial,
TA, tricuspid annulus; ATL, anterior tricuspid leaflet; PTL, posterior tricuspid leaflet; STL, septal tricuspid leaflet, <sup>a</sup>*P* < 0.05 vs mild-moderate, <sup>b</sup>*P*
< 0.05 vs severe.

### Tricuspid Annular Geometry Remodeling

#### Dilatation

The 3D annular area in the massive-torrential group (1642.2 ± 571.3 mm²) was significantly larger than in both the severe (1238.5 ± 296.2 mm²) and mild-moderate (1131.5 ± 261.6 mm²) groups (*P* < 0.001). 3D annular circumference followed the same trend, reaching 144.6 ± 24.0 mm in the massive-torrential group.

#### Planar Flattening

The annulus lost its physiological saddle shape, becoming significantly flatter in the massive-torrential group (annular height: 4.6 ± 1.5 mm) compared to the mild-moderate group (5.5 ± 1.8 mm; *P* = 0.003).

#### Circularization

Analysis revealed a progressive geometric transformation of the TA from an oval to a circular configuration. In the mild-moderate group, the TA shape was generally oval (ellipticity index 1.2 ± 0.3) (Figure 3A). As the disease progressed to the severe stage, the annulus became less elliptical, with an index of 1.1 ± 0.2 (Figure 3B). Notably, in the massive-torrential group, the annulus deformed into a nearly perfect circle, indicated by an ellipticity index of 1.0 ± 0.1 (Figure 3C), which was significantly lower than that of the mild-moderate group (*P* < 0.001).

**Figure 3.**
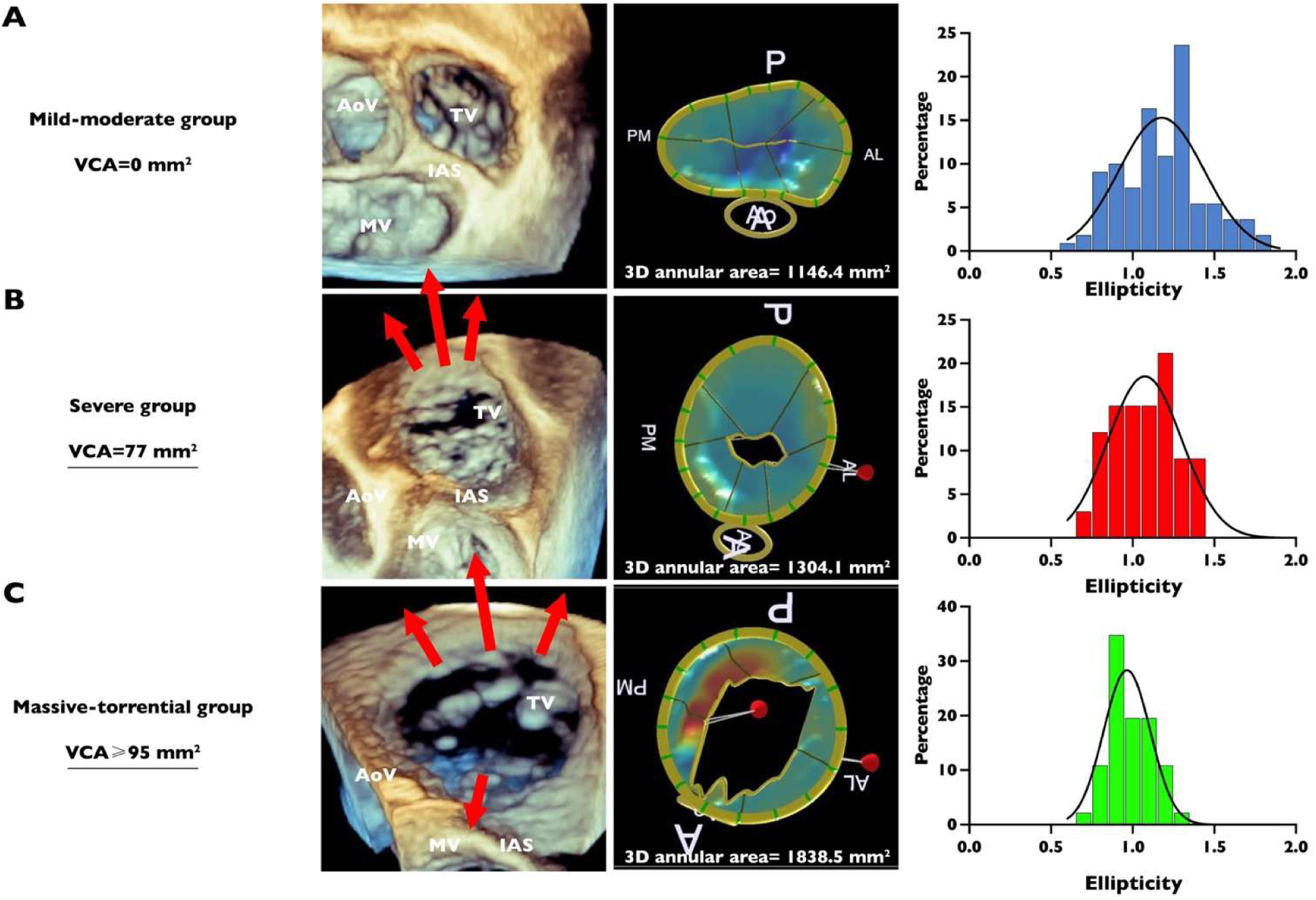
TA shape. Distribution of the ellipticity of TA in mild-moderate(A), severe(B), and massive-torrential (C) FTR patients. A indicates anterior; AL, anterolateral; IAS, interatrial septum; P, posterior; and PM, posteromedial.

#### Orientation

The direction of annular expansion was analyzed to determine the vector of dilatation along the RV free wall (Figure 4A). In the total population, the mean orientation angle (between the LA and the vertical axis at the level of the interatrial septum) was 59.0° ± 23.8°; however, a histogram analysis revealed a bimodal distribution with distinct peaks at approximately 40° and 90° (Figure 4C). When stratified by subgroup, distinct orientation patterns emerged (Figure 4D). The mild-moderate group exhibited a predominantly anterior expansion vector with a mean angle of 40.9° ± 6.2°. In contrast, as illustrated in the representative 3D reconstructions in Figure 4B, the orientation vector shifted significantly in the advanced stages. Both the severe (87.7° ± 11.8°) and massive-torrential (84.7° ± 11.6°) groups displayed significantly larger angles compared to the mild-moderate group (*P* < 0.001), indicating that in advanced FTR, the annulus expands primarily along the anterolateral border.

**Figure 4.**
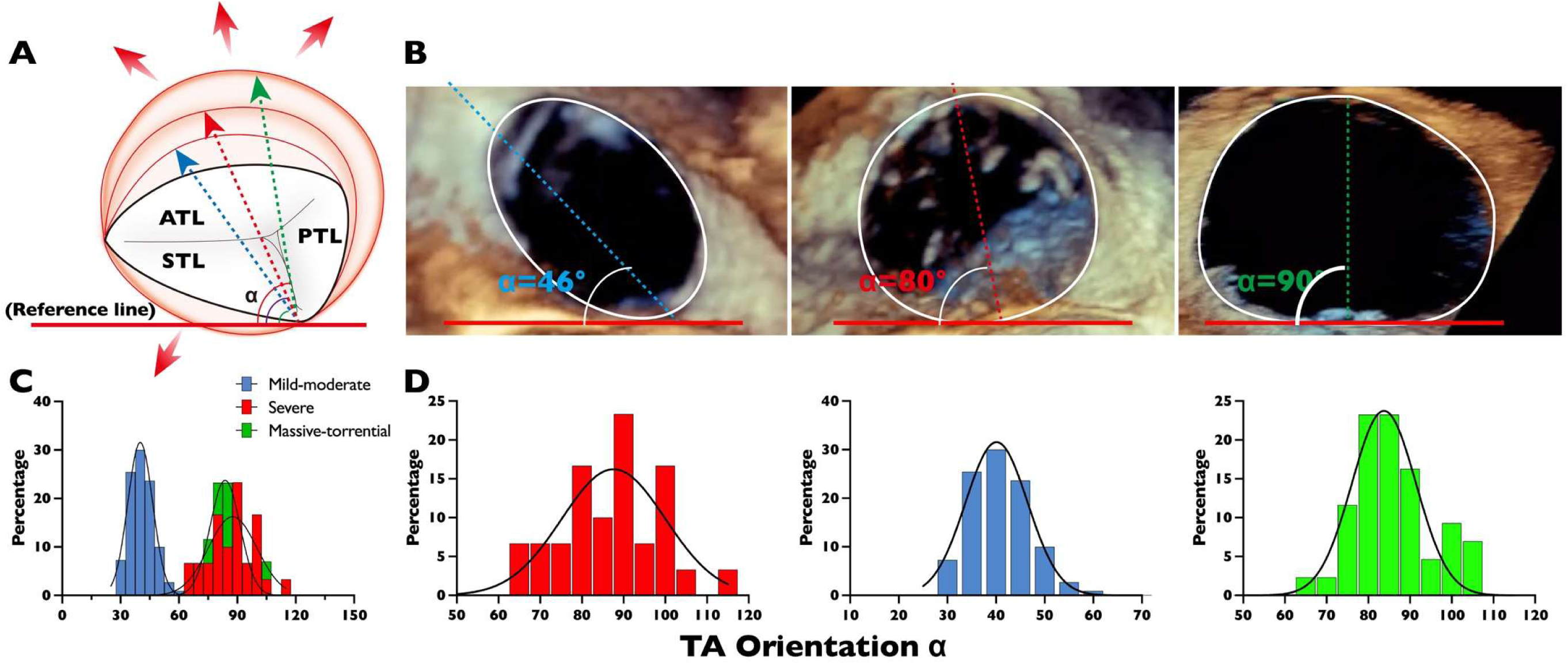
TA orientation. (A), Illustration of progressive TA orientation dilatation. (B), Examples of orientations of the TA. Distribution of the TA orientation of the total FTR patient population (C) and according to subgroups (D). Red arrows show the increasing intercommissural distance with TA dilation. The solid red line represents the reference line parallel to the IAS. The dotted yellow line shows major TA axis. α is the angle between the major TA axis and IAS parallel. A indicates anterior; S, septum; P, posterior; and IAS, interatrial septum.

### Tricuspid Leaflet Remodeling

#### Tethering

The massive-torrential group demonstrated profound tethering, with a tenting volume of 3.2 ± 2.3 mL, significantly exceeding the 1.5 ± 1.2 mL seen in mild-moderate patients (*P* < 0.001). The averaged tethering angle was markedly elevated (42.5° ± 3.0° vs. 15.2° ± 4.4°; *P* < 0.001), reflecting severe apical displacement.

#### Leaflet Morphology

While leaflet thickness significantly increased in the massive-torrential group (*P* < 0.001) (Figure 5), the total leaflet relative length was paradoxically shorter (79.7% ± 28.0%) compared to the mild-moderate group (167.9% ± 39.5%; *P* < 0.001). This indicates a critical uncoupling where leaflet remodeling fails to match the scale of annular dilatation.

**Figure 5.**
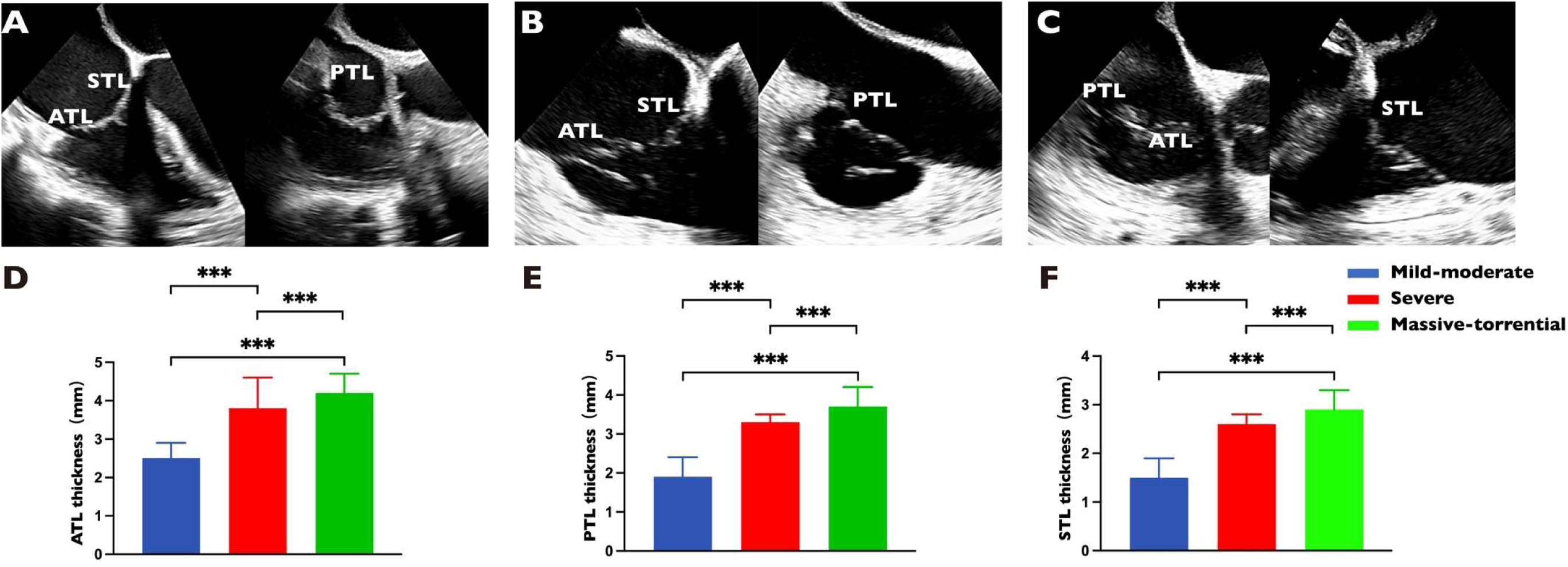
Representative examples of tricuspid leaflet thickness and quantitative analysis. Examples of leaflet thickness in patients with mild-moderate (A), severe (B) and massive-torrential (C) FTR and quantitative analysis of ATL (D), PTL (E) and STL (F). ATL indicates anterior tricuspid leaflet; STL, septal tricuspid leaflet; PTL, posterior tricuspid leaflet.

### Subvalvular Apparatus Remodeling

Quantitative analysis of the subvalvular apparatus revealed significant elongation of the chordae tendineae. The anterior papillary chordae tendineae length increased from 26.1 ± 10.0 mm in the mild-moderate group to 34.0 ± 7.4 mm in the massive-torrential group (*P* < 0.001). Similarly, the septal papillary chordae tendineae length was significantly extended in the massive-torrential group compared to the mild-moderate group (36.4 ± 9.1 mm vs. 31.3 ± 10.3 mm; *P* = 0.009). Additionally, geometric modelling demonstrated significant apical and lateral displacement of the papillary muscles, evidenced by a progressive increase in the subvalvular triangle area (S_△ABC_) and height H in the massive-torrential group (*P* < 0.001) (Table S1).

### Regurgitant Orifice Morphology

The geometry of the regurgitant orifice became increasingly complex with disease progression. While mild-moderate TR typically presented with a lunar-shaped orifice, massive-torrential TR frequently manifested as irregular, non-contiguous shapes (Figure 6A), often displaying two or even three separate orifices within the coaptation line (Figure 6B, C).

**Figure 6.**
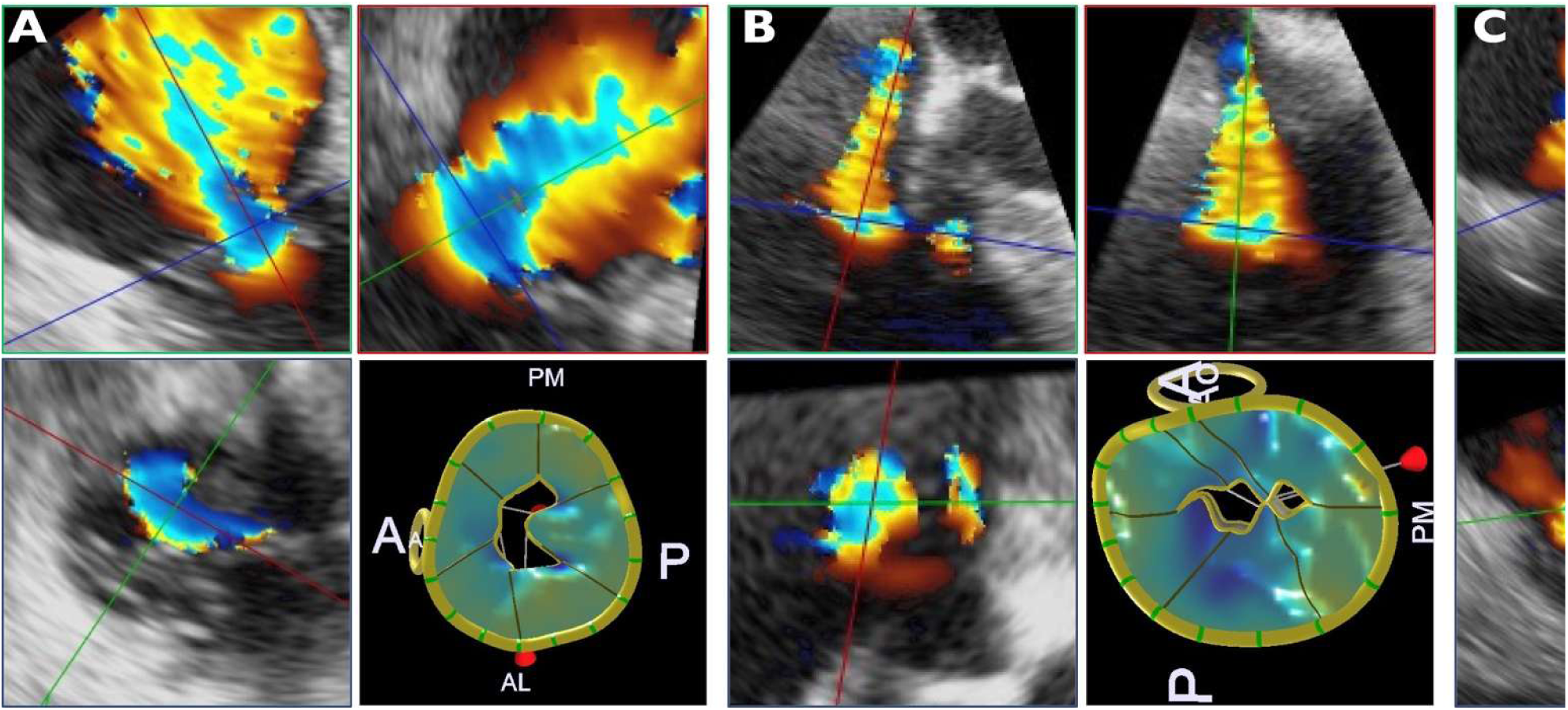
Variable geometry of the FTR orifice. The dataset has been cut transversally to show the FTR orifice from the atrial perspective. the 3D color Doppler image aligns the green and red planes (upper panels) to image the VC in the blue plane (lower panels). Due to the complexity of the tricuspid leaflets, the shape of the TR jet is frequently irregular. In general, the TR jet appears as a lunar cross-section (A), sometimes as two quasi-circular cross-sections (B) or even three quasi-circular cross-section (C). A indicates anterior; AL, anterolateral; IAS, interatrial septum; P, posterior; and PM, posteromedial.

## Discussion

### Major Findings and Clinical Implications

In this study using 3D TEE quantification, we identified a specific “massive/torrential” FTR phenotype characterized by extreme geometric remodeling. Our core findings and their immediate clinical implications are summarized as follows:

1. Distinct geometric phenotype: massive-torrential TR constitutes a distinct structural state defined by extreme annular circularization (ellipticity index ≈ 1.0) and planar flattening, rather than a mere volumetric extension of severe TR. Clinically, this physiologic loss of saddle shape and non-planarity suggests that annuloplasty rings must be sufficiently rigid or 3D-contoured to restore annular geometry and competence;
2. Dual-hit mechanism: these changes are driven by a combined mechanism of RA dilation (leading to annular expansion) and RV remodeling (leading to papillary muscle displacement). This implies that therapeutic strategies addressing a single mechanism may be insufficient for these mixed-etiology patients who also suffer from significant subvalvular tethering; 3) Leaflet-annulus uncoupling: we observed a critical “uncoupling” where compensatory leaflet growth fails to match the scale of massive annular dilation. This mismatch creates large, non-coapting gaps that likely exceed the grasping limits of current edge-to-edge repair devices, necessitating alternative strategies such as leaflet augmentation or replacement; 4) Complex orifice morphology: the regurgitant orifice in massive-torrential grades frequently manifests as complex, multi-focal shapes rather than a simple crescent. This complexity challenges standard 2D VCW grading and underscores the necessity of 3D VCA for accurate risk stratification and procedural planning.

### Mechanisms of Remodeling: The Atrial and Ventricular Interplay

The morphological remodeling observed provides mechanistic validation for the clinical “vicious cycle” of advanced FTR. The high prevalence of AF in the massive–torrential group drives significant RA enlargement, which mechanically pulls the annulus outward ^19^. This “Atrial FTR” mechanism explains the unique circularization and flattening. As the RA dilates, the annulus expands preferentially along the RV free wall, preventing effective leaflet coaptation by spatially separating the leaflet hinge points ^13, 20, 21^.

Simultaneously, the “Ventricular FTR” component is driven by the significant RV dilatation and pulmonary hypertension ^3, 22^. The documented elongation of the chordae tendineae and increased tethering angles reflect the apical displacement of the papillary muscles. This tethering fixes the leaflets in a semi-open position. Crucially, we observed a “maladaptive” leaflet response: although the leaflets were significantly thicker ^23^, their relative length decreased. This “uncoupling” phenomenon results in the large, non-coapting gaps characteristic of torrential disease. The complexity of the regurgitant orifice is a direct consequence of this combined annular distortion and subvalvular tethering ^24^.

### Implications for Transcatheter Interventions

The morphological insights from this study possess direct translational value for the growing field of transcatheter TV interventions. Patients with massive-torrential TR generally exhibit severe leaflet tethering and loss of effective coaptation area, which explains the high failure rate of isolated annuloplasty in such patients. Consequently, these anatomical features suggest that valve replacement may offer a more durable option than repair for this subgroup.

However, the transition to TTVR presents new challenges. Future TTVR strategies may require larger-sized valves to accommodate the extreme annular dilation. Furthermore, attention must be paid to the risk of leaflet perforation or anchoring failure due to the fragile, tethered nature of the leaflets. Anatomical selection criteria from recent pivotal trials, such as TRISCEND II (EVOQUE system) ^25^ and LuX-Valve Plus system ^26^, have highlighted that extreme annular dimensions and severe RV dysfunction remain key exclusion criteria. Our data on “leaflet-annulus uncoupling” provide a morphological basis for current exclusion criteria in pivotal trials, suggesting that pre-procedural 3D TEE screening is critical to identify patients whose anatomy exceeds the limits of current prosthetic valve sizes.

## Limitations

This study was performed at a tertiary center, which may introduce referral bias. Additionally, while we quantified anterior and posterior papillary chordae, the septal papillary muscle was not assessed due to software limitations. Furthermore, hemodynamic assessment via TEE may be subject to altered loading conditions; specifically, in massive or torrential TR, the rapid equalization of RV and right atrial pressures can flatten the TR jet velocity profile, which potentially leads to an underestimation of continuous-wave Doppler-based PASP. Finally, the number of patients in the “torrential” subset specifically was relatively small, warranting larger multicenter studies to confirm these geometric distinctives.

## Conclusions

Massive and torrential FTR are characterized by a unique geometric profile involving extreme annular circularization, severe leaflet tethering, and a critical “leaflet-annulus uncoupling”. These changes are driven by the combined clinical forces of chronic AF and RV remodeling. Understanding this distinct morphology is essential for selecting appropriate interventions, as standard repair techniques may prove insufficient for this complex phenotype.

### Clinical Perspectives

#### Core Clinical Competencies

Massive and torrential FTR represent a distinct morphological phenotype rather than a simple volumetric progression of severe disease. This phenotype is characterized by extreme tricuspid annular circularization (ellipticity index ≈ 1.0) and “leaflet-annulus uncoupling”, where compensatory leaflet growth fails to match massive annular dilation. Clinicians should recognize that these structural changes, often driven by chronic AF and RV remodeling, significantly impact the effectiveness of standard repair techniques.

#### Translational Outlook

The structural complexity of massive-torrential FTR, particularly severe tethering and multi-focal orifices, suggests that isolated annuloplasty carries a high risk of failure in this population. These findings support the consideration of TTVR for these advanced stages, provided that pre-procedural 3D echocardiography confirms anatomical eligibility regarding annular dimensions and RV function. Future research is needed to determine if larger prosthetic valve sizes or leaflet augmentation strategies can overcome the limitations of current “leaflet-annulus uncoupling”.

## Data Availability

The datasets generated and analyzed during the current study are not publicly available due to institutional data privacy and patient confidentiality restrictions but are available from the corresponding author upon reasonable request.

## Acknowledgments

This work was supported by the National Natural Science Foundation of China (Grant No. 82230066, 82001854, 82502385), and the Natural Science Foundation of Hubei Province (Grant No. 2023AFB898).

## Conflict of interest

The authors declare no conflict of interest.

## Notes

### Competing Interest Statement

The authors have declared no competing interest.

### Author Declarations

the Institutional Review Board/Ethics Committee of Union Hospital, Tongji Medical College.

